# Discriminative capacity of the 6-item Vision-related Quality of life and Limitations questionnaire (VQL-6)

**DOI:** 10.1101/2024.10.31.24316475

**Authors:** Vera Linde Dol, Antonius A. J. Roelofs, Anselm B. M. Fuermaier, Anne C. L. Vrijling, Joost Heutink, Nomdo M. Jansonius

## Abstract

**Purpose:** The 6-item Vision-related Quality of life and Limitations questionnaire (VQL-6) is a screening instrument that signals a need for additional low vision care in patients with chronic ophthalmic diseases. This study aims to (1) determine the optimal scoring method for the VQL-6 and (2) evaluate its discriminative capacity for clinical use.

**Methods:** Patients with glaucoma and retina- and cornea-related disorders completed the VQL-6 and participated in an interview designed to distinguish between patients with and without a need of additional care. For the two subscales (general health and quality of life, HQOL, and vision-related limitations, VL) we compared a scoring model with equal item weights to a cross-validated model using regression weights. The optimal scoring method and discriminative capacity of the subscales were determined using receiver operating characteristics (ROC) analysis, with the interview outcome as gold standard. Sensitivities were evaluated at specificities of 90 and 95%.

**Results:** In 297 interview assessments, 96 patients (32%) appeared to need additional care. Both scoring models demonstrated very similar area under the ROC curves (AUC). The selected equal weight model yielded an AUC of 0.91 (95% confidence interval 0.87-0.94) for VL and 0.71 (0.65-0.77) for HQOL. For VL, sensitivity was 72 and 64% at 90 and 95% specificity, respectively. Corresponding HQOL sensitivities were 34 and 17%.

**Conclusions:** The subscale VL shows excellent discriminative capacity and outperformed HQOL. Future studies should explore the feasibility of the VQL-6 in clinical practice.

## Introduction

Visual impairments significantly impact an individual’s quality of life, affecting daily activities and psychological well-being.^1–5^ Additional low vision care, such as prescription and training of visual aids, visual rehabilitation, or medical social work, can improve overall quality of life and daily functioning in people with visual impairments.^6,7^ Yet, it is reported in literature that people are often referred too late to low vision services.^8–12^

While quality of life is of key interest in low vision care settings, the main focus in ophthalmic clinics is on the assessment of visual functions for clinical decision making and referrals. These measures, however, may overlook the nonvisual factors that influence quality of life and rehabilitative needs.^13–15^ Moreover, visual rehabilitation is seen as a last resort by ophthalmologists,^16^ and quality of life is infrequently discussed during consultations.^17^ Consequently, reduced (vision-related) quality of life and a need for additional low vision care in ophthalmic patients may be overlooked, delaying timely referrals.

Questionnaires may be a useful tool to improve patient-clinician communication,^18^ aiding in early detection of reduced quality of life and improving timely referrals.^19,20^ However, existing questionnaires often lack validation for screening purposes or are too time-consuming in clinical practice. To address this gap, we developed the 6-item Vision related Quality of Life and Limitations questionnaire (VQL-6). The VQL-6 is a short questionnaire that measures general health and quality of life, and vision-related limitations, aiming to signal a need for additional low vision care (referred to as ‘additional care’ from hereon) in patients with chronic ophthalmic diseases. In previous work, the VQL-6 demonstrated good psychometric properties in a large Dutch population-based sample and in patients with glaucoma and macular degeneration.^21^ Factor analyses demonstrated a robust two-factor structure, resulting in two subscales: general health and quality of life (HQOL), and vision-related limitations (VL).

The aim of the present study is to (1) determine the optimal scoring method of the VQL-6 and (2) subsequently evaluate its discriminative capacity for clinical use. For this purpose, patients with glaucoma and retina- and cornea-related disorders completed the VQL-6 and took part in a semi-structured interview. The interview was designed to distinguish between patients with and without a need of additional care. We determined the optimal scoring method and the discriminative capacity of the VQL-6 subscales in terms of Receiver Operating Characteristics (ROC) analysis, with the interview outcome as gold standard.

## Methods

### Study population

Participants were consecutive, regular visitors of the outpatient department of Ophthalmology of the University Medical Center Groningen (UMCG) with a scheduled appointment at the cornea, glaucoma, or anti-VEGF injection consultation hour. Patients were eligible to participate if they were 18 years or older, had Dutch as their primary language, and were diagnosed with one or more of the following chronic conditions: glaucoma or retina- or cornea-related disease. The disease had to persist for at least one year and participants had to be under active treatment of an ophthalmologist. We excluded patients who had undergone ophthalmic surgery within the three months preceding their appointment, in order to avoid confounding by temporary changes in visual function.

The ethics board of the UMCG approved the study protocol (#201800249). All participants provided written informed consent. The study followed the tenets of the Declaration of Helsinki.

### Data collection

Eligible patients were asked by the researcher (VLD) to complete the VQL-6 in the waiting room immediately before or after their scheduled appointment. If patients faced challenges due to low vision or other physical limitations, assistance could be offered, e.g., by the person who accompanied the patient. All participants were called by the same researcher within a week’s notice to partake in the interview. The telephone interview took approximately 15 minutes and was recorded with an audio recorder. Responses of the patients, along with additional comments from the researcher, were documented on an interview template. Patient characteristics were collected from their medical files, including age, gender, ophthalmic diagnosis, visual acuity, and perimetry results.

## Materials

### The 6-item Vision-related Quality of life and Limitations (VQL-6)

Details of the VQL-6 have been described before.^21^ The VQL-6 comprises six items (see Supplementary S1 for the Dutch questionnaire; see Supplementary S2 for an English translation). On item 1, 2, and 3, respondents are asked to indicate, on a scale of 0 to 10, their (1) general health, (2) general quality of life, and (3) the extent of experienced limitations by their visual impairment. The subsequent items 4, 5, and 6 measured whether the respondent, because of their visual impairment, (4) worries about the future, (5) feels like he/she accomplishes less, and (6) feels dependent on others. Items 4, 5, and 6 were scored on a 5-point Likert scale with answer options ‘never’, ‘rarely’, ‘sometimes’, ‘often’, and ‘always. The VQL-6 generates one subscale related to general health and quality of life (HQOL; item 1 and item 2), and a vision-specific subscale measuring the extent of vision-related limitations (VL; item 3 to item 6). Respondents were instructed to answer the questions based on their situation in the past month while considering their use of glasses or contact lenses, if applicable. For analysis, all VQL-6 items were converted to a 0 to 10 scale (by transforming the 5-point Likert scale answer options to 0, 2.5, 5, 7.5, 10). In addition, all items were transformed so that a higher score represents better functioning to enable a fair comparison between items.

### Interview

A semi-structured interview with 7 open-ended questions was constructed to assess whether a patient may need additional care and served as a gold standard for the evaluation of the VLQ-6. The first six interview questions were formed by reconstructing the six closed-ended questions of the VQL-6 into open-ended interview questions. Similar to the VQL-6, respondents were instructed to answer the questions based on their situation in the past month. Based on the responses, a comprehensive insight was attained into the general health and quality of life, and the extent of the vision-related limitations experienced by the patients. Interview Question 7 was designed to assess whether the patients themselves indicated a need for additional care. The possibilities (including low vision aids, visual rehabilitation, medical social work, training, and practical advice) were explained to the patients so that they could form an informed opinion. The interview outcome had three scoring options (need for additional care/no need for additional care/uncertain). The three scoring options were later dichotomized into ‘need for additional care’ or ‘no need for additional care’ for analyses, as described below. Scoring of the interview was based on the researcher’s evaluation of the patient’s responses to the first six interview questions. The patient’s self-perceived need for care was also taken into account. For all interview questions, patients were asked to explain their answer and give examples.

To ensure reliability of the interview, we assessed the interrater reliability of the main interview outcome by having a clinical expert (PP, a professional who performs intake interviews at a visual rehabilitation center) score a random sample of 60 interview audio recordings (with scoring options ‘need for additional care’/’no need for additional care’/’uncertain’), covering 20 cornea patients, 20 retina patients, and 20 glaucoma patients. Cases within this sample that were scored differently by the researcher and clinical expert were discussed until a consensus was reached. In addition, to dichotomize the main interview outcome into ‘need for additional care’ or ‘no need for additional care’, audio recordings of all cases of the total sample that were categorized by the researcher as ‘uncertain’ on Question 7 were discussed between the researcher and clinical expert and reevaluated.

### Data analysis

Data analysis was performed using RStudio (version 2023.12.0).^22^ Data was checked for normality and linearity; if the assumptions were violated, non-parametric tests were performed. A p-value of 0.05 or less was considered statistically significant.

### Descriptives

To evaluate results across different ophthalmic conditions, all data was categorized into four ophthalmic conditions: retina (e.g., macular degeneration, diabetic retinopathy, or retinal vein occlusion), glaucoma, cornea (e.g., keratoconus, corneal infection, corneal dystrophy), and comorbid. The comorbid category included patients with a combined retina, cornea, or glaucoma diagnosis. We did not consider other ocular comorbidities, such as cataract, amblyopia or myopia, in our categorization, that is, for example, a glaucoma patient with myopia was classified in the glaucoma category. The participant characteristics were described with mean and standard deviation (SD). For patients with visual field test results available (the glaucoma patients, including those within the comorbid group), the integrated visual field (IVF) score was calculated from the monocular visual field test results extracted from the HFA printouts, using the ‘best sensitivity’ method.^23,24^ Age, gender, main interview outcomes, and VQL-6 subscale sum scores were compared between the patient groups using Kruskal-Wallis test and chi-square (χ^2^) tests.

For the interview, descriptive data was generated for the interview outcome. Cohen’s kappa with linear weighting was established as a measure of interrater reliability of the interview (applied to the 3×3 table with the options ‘need for care’/’no need for care’/’uncertain’). Values < 0 indicate agreement worse than chance, 0–0.20 none to slight, 0.21–0.40 fair, 0.41–0.60 moderate, 0.61–0.80 substantial, and 0.81–1.00 almost perfect agreement.^25^ VQL-6 subscale scores between participants with and without a need for additional care based on the interview were compared with Wilcoxon rank sum tests.

### Scoring method and discriminative capacity

To determine the optimal scoring method of the VQL-6 subscales, we compared two models. First, we evaluated an equal weight model, which was determined on the a priori assumption that all items have equal weight in predicting a need for additional care, signifying simple sum scores for each subscale. Second, we evaluated a cross-validated fitted model - a competing model that was fitted to our dataset - where each item was weighted according to its contribution in predicting a need for additional care. For the fitted model, we first performed a binary logistic regression for both subscales, with the subscale item scores as independent variables and the interview outcome as a dependent variable. Weighted subscale scores were calculated using the Beta values of the regression formula giving weight to each item. ROC analyses were performed for the equal weight model and the fitted model, with the interview outcome as gold standard, employing the pROC package in R.^26^ Considering the fitted model was built and tested in the same dataset, we performed a 10-fold cross validation using the ‘caret’ package in R,^27^ to estimate how well the ROC model would perform in an independent dataset^28^ Mean and standard deviation of the 10 evaluations represented the generalized performance and standard error of the fitted model.

To evaluate which model performed best, the AUC values of the equal weight model and the fitted model were compared for significant differences, using bootstrap tests for correlated AUC values.^29^ Within each model, we determined the best performing VQL-6 subscale by comparing AUC values employing DeLong’s test.^30,31^ AUC values were interpreted according to the following criteria: AUC < 0.7 low, 0.7 to 0.9 acceptable, and ≥ 0.90 excellent.^32^ Using the best performing model, sensitivities were evaluated at predefined specificity values of 90% and 95%.

Finally, we performed ROC analyses to evaluate the capacity of visual functions in predicting a need for additional care. AUC values were calculated and compared with the VQL-6 subscales, for visual acuity (in logMAR) of the better eye for all patients and for HFA MD of the better eye and the integrated visual field (IVF) for the glaucoma patients and comorbid patients with glaucoma.

## Results

### Descriptives

Table 1 presents the participant characteristics. In total, 322 patients agreed to participate. One participant was excluded based on missing VQL-6 responses. In addition, 24 participants were excluded as they did not complete the interview for various reasons, including no registered phone number, did not pick up the phone, or not being able to partake due to bad hearing or poor cognitive capabilities. After exclusion, 297 participants were left for data analysis. Included patients were categorized according to their ophthalmic diagnosis, resulting in 96 retina, 60 glaucoma, 81 cornea, and 60 comorbid patients. There was a significant difference between patient groups with respect to age (*p* <.001). No significant difference was found for gender (*p* = 0.51).

**Table 1.**
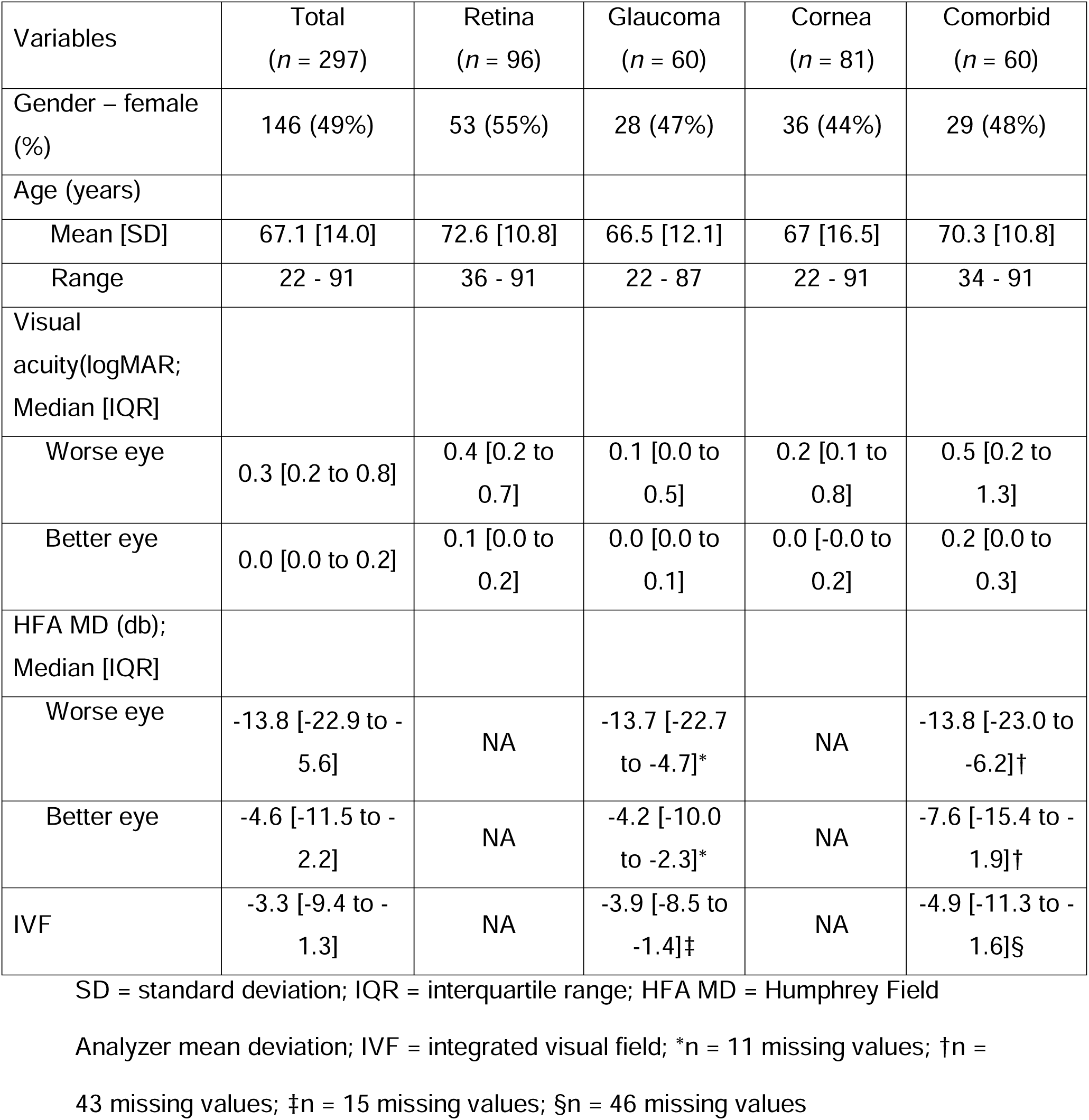
Characteristics of the study population.

| Variables | Total<br>(*n* = 297) | Retina<br>(*n* = 96) | Glaucoma<br>(*n* = 60) | Cornea<br>(*n* = 81) | Comorbid<br>(*n* = 60) |
| --- | --- | --- | --- | --- | --- |
| Gender – female (%) | 146 (49%) | 53 (55%) | 28 (47%) | 36 (44%) | 29 (48%) |
| Age (years) |  |  |  |  |  |
| Mean [SD] | 67.1 [14.0] | 72.6 [10.8] | 66.5 [12.1] | 67 [16.5] | 70.3 [10.8] |
| Range | 22 - 91 | 36 - 91 | 22 - 87 | 22 - 91 | 34 - 91 |
| Visual acuity(logMAR; Median [IQR]) |  |  |  |  |  |
| Worse eye | 0.3 [0.2 to 0.8] | 0.4 [0.2 to 0.7] | 0.1 [0.0 to 0.5] | 0.2 [0.1 to 0.8] | 0.5 [0.2 to 1.3] |
| Better eye | 0.0 [0.0 to 0.2] | 0.1 [0.0 to 0.2] | 0.0 [0.0 to 0.1] | 0.0 [-0.0 to 0.2] | 0.2 [0.0 to 0.3] |
| HFA MD (db); Median [IQR] |  |  |  |  |  |
| Worse eye | -13.8 [-22.9 to -5.6] | NA | -13.7 [-22.7 to -4.7]* | NA | -13.8 [-23.0 to -6.2]† |
| Better eye | -4.6 [-11.5 to -2.2] | NA | -4.2 [-10.0 to -2.3]* | NA | -7.6 [-15.4 to -1.9]† |
| IVF | -3.3 [-9.4 to -1.3] | NA | -3.9 [-8.5 to -1.4]‡ | NA | -4.9 [-11.3 to -1.6]§ |

SD = standard deviation; IQR = interquartile range; HFA MD = Humphrey Field
Analyzer mean deviation; IVF = integrated visual field; \**n* = 11 missing values; †*n* =
43 missing values; ‡*n* = 15 missing values; §*n* = 46 missing values

Table 2 shows the median sum scores of the VQL-6 subscales, stratified per interview outcome and per patient group. Of all 297 patients, 96 (32%) were evaluated to be in need of additional care based on the interview. Using the data from Table 2 multiple comparisons were made, both between interview outcomes (horizontally) and between different patient groups (vertically). Significant differences were found for both subscales per interview outcome, with patients with a need for additional care scoring significantly lower on HQOL and VL than patients without a need for additional care (both *p* < 0.001). This was the case for all patients together and for the individual patient groups separately (all *p* < 0.001). Between the patient groups, no significant differences were found in the HQOL scores (*p* = 0.23), but a significant difference was found in the VL scores (*p* = 0.008). Post-hoc Dunn tests demonstrated significantly lower VL scores for cornea patients, compared to glaucoma (*p* = 0.04) and retina (*p* = 0.04) patients. Additionally, no significant differences were found in the main interview outcome between the patient groups (*p* = 0.33). Therefore, the patient groups were merged, and all additional analyses were performed on the group as a whole.

**Table 2.**
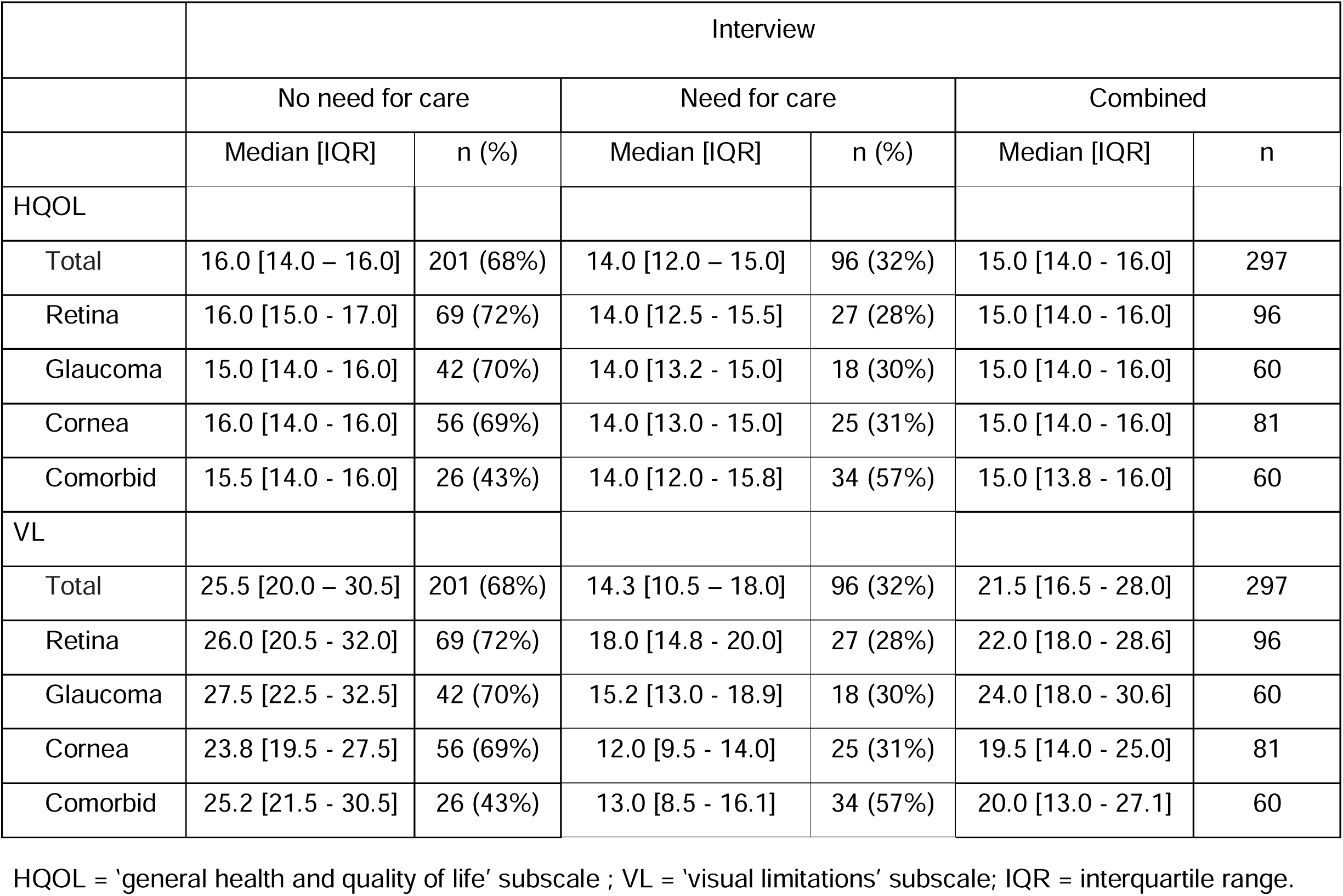
Subscale sum scores of patients stratified per interview outcome and per patient group.

### Scoring method and discriminative capacity

In our total sample, the binary logistic regression analysis of HQOL and VL resulted in the following classification equations:

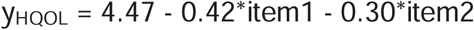

and

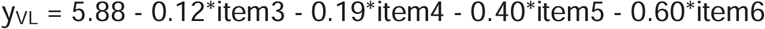

Weighted subscale scores were calculated and a 10-fold cross validation was performed to calculate the generalized performance of the fitted model. Figure 1 shows the ROC curves and AUC values of the equal weight model and the fitted model, for both subscales. For comparison, we added an ROC curve and AUC value for the visual acuity of the better eye. HQOL showed acceptable discriminative capacity in both models (AUC equal weight [95% CI] = 0.71 [0.65 - 0.77]; AUC fitted [95% CI] = 0.71 [0.65 - 0.77]). VL demonstrated excellent discriminative capacity (AUC equal weight [95% CI] = 0.91 [0.87 - 0.94]; AUC fitted [95% CI] = 0.92 [0.89 - 0.95]). A comparison between subscales demonstrated a significantly higher AUC value of VL compared to HQOL, using the equal weight model (*Z* = 6.45, *p* < .001) and the fitted model (*Z* = 6.86, *p* < .001). Between the equal weight and the fitted model, no significant differences were found for HQOL (*D* = 0.13, *p* = 0.90) and VL (*D* = 0.46, *p* = 0.65). Therefore, following parsimony, we employed the equal weight model for the evaluation of sensitivity and specificity, and the subsequent analysis.

**Figure 1.**
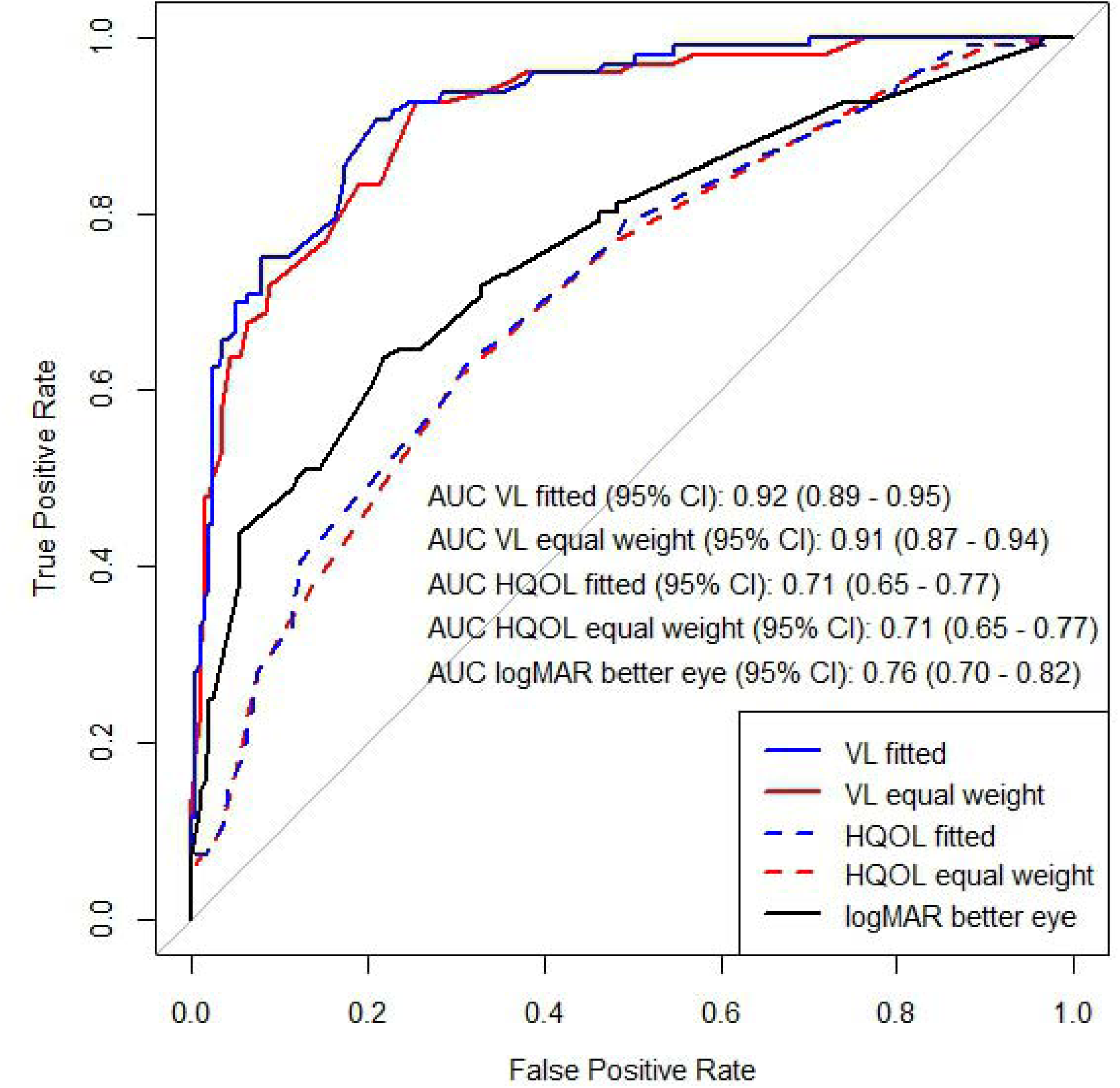
Receiver operating characteristics curves of the VQL-6 subscales (VL, vision-related limitations; HQOL, general health and quality of life) using the equal weight model and the fitted model and visual acuity (logMAR better eye) in discriminating between patients with ‘a need for additional care’ versus ‘no need for additional care’. AUC = area under the curve.

Table 3 shows sensitivities and specificities (specificity starting from 70%) for potential cutoff sum scores of the VL subscale. At a predefined specificity level closest to 90% (91%), a sensitivity of 72% was achieved, with a cutoff sum score of ≤ 17.75. At a specificity level closest to 95% (95.5%), a sensitivity of 64% was found, with a cutoff sum score of ≤ 15.75. For HQOL sensitivity levels of 34% and 17% were found for specificities of 90 or 95%, respectively. Given this poor performance of HQOL, we did not tabulate potential cutoff scores.

**Table 3.** Summary of cutoff scores of the VL subscale.

| Cutoff | Sensitivity | Specificity |
| --- | --- | --- |
| $\leq 21.25$ | 92.7% | 71.6% |
| $\leq 20.75$ | 92.7% | 73.1% |
| $\leq 20.25$ | 92.7% | 74.6% |
| $\leq 19.75$ | 83.3% | 78.6% |
| $\leq 19.25$ | 83.3% | 81.1% |
| $\leq 18.75$ | 79.2% | 83.6% |
| $\leq 18.25$ | 77.1% | 84.6% |
| $\leq 17.75$ | 71.9% | 91.0% |
| $\leq 17.25$ | 68.8% | 91.5% |
| $\leq 16.75$ | 67.7% | 93.5% |
| $\leq 16.25$ | 63.5% | 94.5% |
| $\leq 15.75$ | 63.5% | 95.5% |
| $\leq 15.25$ | 58.3% | 96.5% |
| $\leq 14.75$ | 53.1% | 96.5% |
| $\leq 14.25$ | 50.0% | 97.5% |

As shown in Figure 1, logMAR (of the better eye) demonstrated an AUC of 0.76 (95% CI: 0.70 - 0.82). VL demonstrated a significantly higher AUC value (*D* = - 4.07, *p* < .001) compared to logMAR. No significant difference was found between logMAR and the AUC value HQOL (*D* = 1.32, *p* = 0.19). Figure 2 depicts the ROC curves and AUC values of both subscales, compared to HFA MD of the better eye and the integrated visual field (IVF) for the glaucoma patients and comorbid patients with glaucoma. HFA MD (of the better eye) yielded an AUC of 0.77 (95% CI: 0.63 - 0.90), which was not significantly different from HQOL (Z = 0.79, p = 0.43) and significantly lower than VL (*Z* = −2.50, *p* = 0.01). Integrated visual field (IVF) measures demonstrated an AUC of 0.81 (95% CI: 0.68 - 0.95), with no significant difference compared to HQOL (*Z* = 1.75, *p* = 0.08) and VL (*Z* = −1.95, *p* = 0.052).

**Figure 2.**
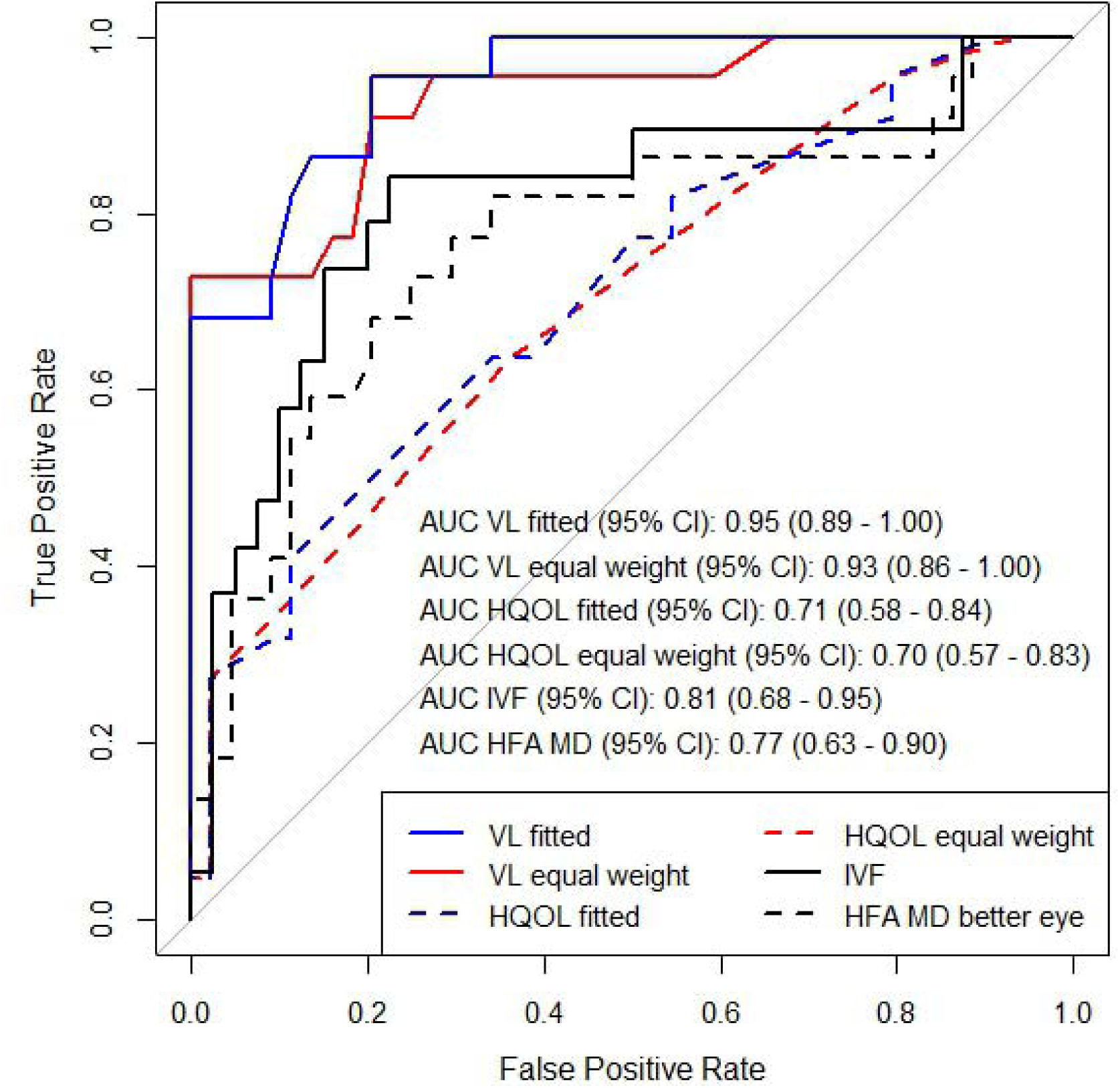
Receiver operating characteristics curves of the VQL-6 subscales (VL, vision-related limitations; HQOL, general health and quality of life) using the equal weight model and the fitted model, the Humphrey Field Analyzer mean deviation (HFA MD better eye), and integrated visual field (IVF) in discriminating between glaucoma patients with ‘a need for additional care’ versus ‘no need for additional care’. AUC = area under the curve.

## Discussion

This study determined the optimal scoring method and discriminative capacity of the VQL-6 subscales in patients with chronic ophthalmic diseases. Through interview assessments, 32% of the patients were evaluated to be in need of additional care. A scoring model with equal weight items and a cross-validated fitted model using regression weights demonstrated a very similar AUC for predicting a need for additional care. Following parsimony, we selected the equal weight model, and the VL subscale showed excellent capacity in predicting a need for additional care. VL outperformed the HQOL subscale, which showed acceptable discriminative capacity, and visual acuity and visual field measures, which revealed good discriminative capacity.

We opted for a scoring method employing equal weighted items instead of weighted items for calculating subscale scores, as both methods resulted in a very similar discriminative capacity of the VQL-6. These results agree with prior studies that have compared equal weighted sum scores with weighted scores derived from item response theory methods or factor analyses.^33–35^ Similarly, studies have demonstrated a comparable psychometric basis of sum scores compared to weighted scores, when the dimensional structure of items is verified.^33,36,37^

We evaluated sensitivity at clinically meaningful specificity levels of 90% and 95%. A specificity lower than these levels would result in a larger number of false positives, which would unnecessarily burden scarce health care resources. HQOL did not reach sufficient levels of combined sensitivity and specificity. For VL, a specificity of 91% (the nearest possible value to 90%), resulted in sufficient sensitivity of 72%. A specificity of 95% resulted in a significant drop of sensitivity to 64%, meaning a significant proportion of patients that may need additional care would be missed, which is not deemed preferable for a screening instrument. In the specific setting of our study, a tertiary referral clinic, a cutoff at a specificity of 91%, would imply a positive predictive value of 79% and a negative predictive value of 87%. Presumably, these numbers may be different in a general hospital, where the prevalence of visual impairment and experienced vision-related limitations might be lower than in our setting - in such a setting it might be better to opt for a higher specificity.

To our knowledge, sensitivity and specificity of other questionnaires in predicting a need for additional care have not been reported. Whilst some studies reported the sensitivity and specificity of questionnaires in predicting measurements of visual functions or discriminating between patients and controls,^38,39^ due to the use of different reference standards, direct comparison with our results is not possible. Guo et al developed an Electronic Medical Record (EMR)-based tool for ophthalmic patients in need of referral to low vision services that fires an alert based on visual acuity and diagnosis criteria.^40^ They reported sensitivity and specificity based on when the firing of the alert should have been suppressed or when firing criteria were not met,^40^ which limits direct comparison to our results. Furthermore, the use of visual acuity-based criteria as a reference standard may not be optimal, as our study’s findings confirm that visual acuity is less accurate in predicting a need for additional care.

Our study revealed the highest capacity of self-reported vision-related limitations in predicting a need for additional care, measured by the subscale VL. Visual acuity was an important predictor as well, yet it was inferior to VL. This is similar to prior research that showed that various factors next to visual acuity were significant predictors of rehabilitation needs, including physical and mental health, comorbidity and dependence of others.^6,13^ While some studies, including O’Connor et al,^13^ have identified visual acuity as the most important predictor (contrasting with our findings), others did not find a significant association between the prevalence of rehabilitation needs and visual acuity.^14,42^ Taken together, referral practices should not be guided by measures of visual function in isolation. The importance of considering other factors, such as emotional wellbeing and comorbidities, beyond visual acuity is clear. This could be achieved by asking the appropriate questions through quality of life questionnaires,^43^ for which the VQL-6 may offer a practical solution.

Our study presents both strengths and limitations. A strength of this study is the evaluation of sensitivity and specificity of our questionnaire. In the absence of an established gold standard, we employed a carefully constructed interview.^44^ Although we found acceptable interrater reliability, a note of caution is due here as the outcome of the interview is inevitably of subjective nature and may be biased by our own personal concept of needing additional care. Moreover, as the interview is partly based on the VQL-6, the resultant upward bias may lead to an overstatement of the discriminative capacity of the VQL-6. All considered, further exploration of the diagnostic capacity of the VQL-6 is warranted. Future research could evaluate the accuracy of referrals guided by the VQL-6, by investigating the outcomes and effectiveness of the referred patients at their follow-up in integrated care. Another strength of this study is the inclusion of a variety of chronic ophthalmic diseases that may benefit from additional care, including patients with comorbidity of (chronic) ophthalmic diseases. Most studies exclude comorbidities or validate questionnaires in specific patient groups, limiting external validity. Yet, a limitation is that all patients were included from a single university medical center study. Therefore, generalizability of our results on the discriminative capacity of the VQL-6 and the suggestions for cut-off criteria to other ophthalmic patient populations in different settings is not self-evident.

The VQL-6 offers a standardized and automated method to screen for patients in need of additional care. A cutoff of ≤ 17.75 on the VL subscale showed good sensitivity and specificity, and may be an adequate criterion for use in clinical practice. It is important to recognize that a VQL-6 alert does not imply a direct referral, as patients may be familiar with low vision services, have an active medical plan, or the visual impairment is expected to improve. It is up to the ophthalmologist to consider all factors and determine - together with the patient - if a referral to additional care is necessary. We found no significant differences in interview outcomes and VQL-6 subscale scores between different patient groups, aligning with previous studies that found that ophthalmic diagnosis did not have a significant effect on quality of life outcomes or rehabilitation needs.^6,13,45,46^ This suggests that the VQL-6 is equally applicable across ophthalmic conditions. Administration of the VQL-6 in EMR ahead of consultations may offer an effective method and minimizes time constraints.^47^ As the VQL-6 is short and a simple sum score is employed, paper administration provides an alternative for those without digital access.

In conclusion, the VQL-6 appears to be a valid instrument with good discriminative capacity. For clinical practice, a cutoff score of 17.75 and lower on the VL subscale may be a good criterion to screen for patients in need of additional care. Future studies should explore the use of the VQL-6 in a clinical setting and evaluate its feasibility in reducing late referrals to low vision rehabilitation.

## Data Availability

The participants of this study did not give written consent for their data to be shared publicly, so due to the sensitive nature of the research supporting data is not available.

## Acknowledgments

We thank Petra Pijnakker for her valuable contribution as a clinical expert to the reassessments of the interviews.

## References

1. Haymes SA, Johnston AW, Heyes AD. Relationship between vision impairment and ability to perform activities of daily living. Ophthalmic Physiol Opt. 2002;22(2):79–91.

2. Bibby SA, Maslin ER, McIlraith R, Soong GP. Vision and selfLreported mobility performance in patients with low vision. Clin Exp Optom. 2007;90(2):115–123.

3. Wood JM, Lacherez P, Black AA, Cole MH, Boon MY, Kerr GK. Risk of falls, injurious falls, and other injuries resulting from visual impairment among older adults with age-related macular degeneration. Invest Ophthalmol Vis Sci. 2011;52(8):5088–5092.

4. Zheng DD, Swenor BK, Christ SL, West SK, Lam BL, Lee DJ. Longitudinal Associations Between Visual Impairment and Cognitive Functioning: The Salisbury Eye Evaluation Study. JAMA Ophthalmol. 2018;136(9):989–995.

5. Evans JR, Fletcher AE, Wormald RPL. Depression and anxiety in visually impaired older people. Ophthalmology. 2007;114(2):283–288.

6. Lamoureux EL, Hassell JB, Keeffe JE. The determinants of participation in activities of daily living in people with impaired vision. Am J Ophthalmol. 2004;137(2):265–270.

7. Gothwal VK, Bharani S. Outcomes of Multidisciplinary Low Vision Rehabilitation in Adults. Investigative Opthalmology & Visual Science. 2015;56(12):7451.

8. Goldstein JE, Guo X, Boland M V, Swenor BK. Low Vision Care - Out of Site. Out of Mind. Ophthalmic Epidemiol. 2020;27(4):252–258.

9. Chiang PPC, O’Connor PM, Le Mesurier RT, Keeffe JE. A Global Survey of Low Vision Service Provision. Ophthalmic Epidemiol. 2011;18(3):109–121.

10. Coker MA, Huisingh CE, McGwin G, et al. Rehabilitation Referral for Patients With Irreversible Vision Impairment Seen in a Public Safety-Net Eye Clinic. JAMA Ophthalmol. 2018;136(4):400–408.

11. Kumar H, Monira S, Rao A. Causes of Missed Referrals to Low-Vision Rehabilitation Services: Causes in a Tertiary Eye Care Setting. Semin Ophthalmol. 2016;31(5):452–458.

12. Pollard TL, Simpson JA, Lamoureux EL, Keeffe JE. Barriers to accessing low vision services. Ophthalmic Physiol Opt. 2003;23(4):321–327.

13. O’Connor PM, Lamoureux EL, Keeffe JE. Predicting the need for low vision rehabilitation services. Br J Physiol Opt. 2008;92(2):252–255.

14. Macnaughton J, Latham K, VianyaLEstopa M. Rehabilitation needs and activity limitations of adults with a visual impairment entering a low vision rehabilitation service in England. Ophthalmic Physiol Opts. 2019;39(2):113–126.

15. Trillo AH, Dickinson CM. The impact of visual and nonvisual factors on quality of life and adaptation in adults with visual impairment. Invest Ophthalmol Vis Sci. 2012;53(7):4234–4241. doi: 10.1167/iovs.12-9580

16. Overbury O, Wittich W. Barriers to low vision rehabilitation: the Montreal Barriers Study. Invest Ophthalmol Vis Sci. 2011;52(12):8933–8938.

17. Sleath B, Sayner R, Vitko M, et al. Glaucoma patient-provider communication about vision quality-of-life. Patient Educ Couns. 2017;100(4):703–709.

18. Greenhalgh J, Gooding K, Gibbons E, et al. How do patient reported outcome measures (PROMs) support clinician-patient communication and patient care? A realist synthesis. J Patient Rep Outcomes. 2018;2(1):42.

19. Dean S, Mathers JM, Calvert M, et al. “The patient is speaking”: discovering the patient voice in ophthalmology. Br J Physiol Opt. 2017;101(6):700–708.

20. Robertson AO, Tadić V, Rahi JS. Attitudes, experiences, and preferences of ophthalmic professionals regarding routine use of patient-reported outcome measures in clinical practice. PLoS One. 2020;15(12):e0243563.

21. Dol VL, Fuermaier ABM, Roelofs TAJ, Vrijling ACL, Heutink J, Jansonius NM. The 6-Item Vision-Related Quality of Life and Limitations Questionnaire: Evaluation of Psychometric Properties. Transl Vis Sci Technol. 2024;13(3):5.

22. RStudio Team. RStudio: integrated Development for R. Published online 2022. Accessed March 20, 2023. http://www.rstudio.com/

23. Asaoka R, Crabb DP, Yamashita T, Russell RA, Wang YX, Garway-Heath DF. Patients Have Two Eyes!: Binocular versus Better Eye Visual Field Indices. Invest Ophthalmol Vis Sci. 2011;52(9):7007.

24. Crabb DP, Viswanathan AC. Integrated visual fields: a new approach to measuring the binocular field of view and visual disability. Graefes Arch Clin Exp Ophthalmol. 2005;243(3):210–216.

25. Cohen J. Weighted kappa: Nominal scale agreement provision for scaled disagreement or partial credit. Psychol Bull. 1968;70(4):213–220.

26. Robin X, Turck N, Hainard A, et al. pROC: an open-source package for R and S+ to analyze and compare ROC curves. BMC Bioinformatics. 2011;12(1):77.

27. Kuhn M. Building Predictive Models in R Using the caret Package. 2008;28:1–26. doi:10.18637/jss.v028.i05

28. Steyerberg EW. Validation of Prediction Models. Clinical Prediction Models. Statistics for Biology and Health. New York, Springer, 2009: 299–311.

29. Carpenter J, Bithell J. Bootstrap confidence intervals: When, which, what? A practical guide for medical statisticians. Stat Med. 2000;19(9):1141–1164.

30. DeLong ER, DeLong DM, Clarke-Pearson DL. Comparing the Areas under Two or More Correlated Receiver Operating Characteristic Curves: A Nonparametric Approach. Biometrics. 1988;44(3):837.

31. Sun X, Xu W. Fast Implementation of DeLong’s Algorithm for Comparing the Areas Under Correlated Receiver Operating Characteristic Curves. IEEE Signal Process Lett. 2014;21(11):1389–1393.

32. Streiner DL, Cairney J. What’s under the ROC? An Introduction to Receiver Operating Characteristics Curves. Can J Psychiatry. 2007;52(2):121–128.

33. Bobko P, Roth PL, Buster MA. The Usefulness of Unit Weights in Creating Composite Scores. Organ Res Methods. 2007;10(4):689–709.

34. Dougherty BE, Bullimore MA. Comparison of scoring approaches for the NEI VFQ-25 in low vision. Optom Vis Sci. 2010;87(8):543–548.

35. Goldstein JE, Bradley C, Gross AL, Jackson M, Bressler N, Massof RW. The NEI VFQ-25C: Calibrating Items in the National Eye Institute Visual Function Questionnaire-25 to Enable Comparison of Outcome Measures. Transl Vis Sci Technol. 2022;11(5):10–10.

36. Widaman KF, Revelle W. Thinking thrice about sum scores, and then some more about measurement and analysis. Behav Res Methods. 2022;55(2):788–806.

37. Ferrando PJ, Chico E. The External Validity of Scores Based on the Two-Parameter Logistic Model: Some Comparisons between IRT and CTT. Psicologica. 2007;28(2):23–257.

38. Coren S, Hakstian AR. Visual screening without the use of technical equipment: preliminary development of a behaviorally validated questionnaire. Appl Opt. 1987;26(8):1468–1472.

39. Wolffsohn JS, Jackson J, Hunt OA, et al. An enhanced functional ability questionnaire (faVIQ) to measure the impact of rehabilitation services on the visually impaired. Int J Ophthalmol. 2014;7(1):77–85.

40. Goldstein JE, Guo X, Swenor BK, Boland M V., Smith K. Using Electronic Clinical Decision Support to Examine Vision Rehabilitation Referrals and Practice Guidelines in Ophthalmology. Transl Vis Sci Technol. 2022;11(10):8.

41. Guo X, Swenor BK, Smith K, Boland M V., Goldstein JE. Developing an Ophthalmology Clinical Decision Support System to Identify Patients for Low Vision Rehabilitation. Transl Vis Sci Technol. 2021;10(3):24.

42. Brown JC, Goldstein JE, Chan TL, Massof R, Ramulu P, Low Vision Research Network Study Group. Characterizing functional complaints in patients seeking outpatient low-vision services in the United States. Ophthalmology. 2014;121(8):1655–62.e1.

43. Luu W, Kalloniatis M, Bartley E, et al. A holistic model of low vision care for improving visionLrelated quality of life. Clin Exp Optom. 2020;103(6):733–741.

44. Rutjes A, Reitsma J, Coomarasamy A, Khan K, Bossuyt P. Evaluation of diagnostic tests when there is no gold standard. A review of methods. Health Technol Assess (Rockv). 2007;11(50).

45. Evans K, Law SK, Walt J, Buchholz P, Hansen J. The quality of life impact of peripheral versus central vision loss with a focus on glaucoma versus age-related macular degeneration. Clin Ophthalmol. 2009;3(1):433–445.

46. Ugurlu SK, Altundal AEK, Ekin MA. Comparison of vision-related quality of life in primary open-angle glaucoma and dry-type age-related macular degeneration. Eye (Lond). 2017;31(3):395–405.

47. Nelson EC, Eftimovska E, Lind C, Hager A, Wasson JH, Lindblad S. Patient reported outcome measures in practice. BMJ. 2015;350:g7818.

