## Supplementary S1 for "Discriminative capacity of the 6-item Vision-related Quality of life and Limitations questionnaire (VQL-6)"

### Instructie

**Lees eerst de vraag en de antwoordmogelijkheden.** Kruis daarna het juiste antwoord aan. Het is belangrijk dat u uw eigen mening geeft en de vragen zonder overleg met anderen invult.

**Er is per vraag maar 1 antwoord mogelijk.** Vul de hele vragenlijst in en sla alstublieft geen vragen over! Wanneer u het antwoord niet precies weet mag u een schatting geven.

**U heeft een fout gemaakt?** Als u een fout maakt is dat niet erg. Maak het foute antwoord in zijn geheel zwart en kruis het juiste antwoord aan. Oftewel:

■ = fout antwoord
☒ = juist antwoord

**Beantwoord de vragen alsof u uw bril of contactlenzen draagt**, als u die heeft.

De vragen in deze vragenlijst hebben betrekking op **uw situatie in de afgelopen maand.**

Dol VL, Fuermaier ABM, Roelofs TAJ, Vrijling ACL, Heutink J, Jansonius NM. The 6-Item Vision-Related Quality of Life and Limitations Questionnaire: Evaluation of Psychometric Properties. *Transl Vis Sci Technol*. 2024;13(3):5. doi:10.1167/tvst.13.3.5. CC BY-NC-SA 4.0

1. Hoe waardeert u uw gezondheid uitgedrukt in een rapportcijfer?

□ 0 □ 1 □ 2 □ 3 □ 4 □ 5 □ 6 □ 7 □ 8 □ 9 □ 10

2. Hoe waardeert u uw kwaliteit van leven uitgedrukt in een rapportcijfer?

□ 0 □ 1 □ 2 □ 3 □ 4 □ 5 □ 6 □ 7 □ 8 □ 9 □ 10

3. In hoeverre wordt u in uw dagelijks leven gehinderd door problemen met uw zicht? (*0 = geen hinder, 10 = zeer ernstige hinder*)

□ 0 □ 1 □ 2 □ 3 □ 4 □ 5 □ 6 □ 7 □ 8 □ 9 □ 10

4. Maakt u zich zorgen over de toekomst vanwege problemen met het zien?

□ nooit
□ zelden
□ soms
□ vaak
□ altijd

**Zie vraag 5 en 6 op de volgende pagina**

Dol VL, Fuermaier ABM, Roelofs TAJ, Vrijling ACL, Heutink J, Jansonius NM. The 6-Item Vision-Related Quality of Life and Limitations Questionnaire: Evaluation of Psychometric Properties. *Transl Vis Sci Technol*. 2024;13(3):5. doi:10.1167/tvst.13.3.5. CC BY-NC-SA 4.0

5. Krijgt u vanwege het zien minder voor elkaar dan u zou willen of kunt u dingen minder goed volhouden? (Denk aan lezen, werken, huishoudelijke taken, eropuit gaan of het beoefenen van hobby's)

□ nooit
□ zelden
□ soms
□ vaak
□ altijd

6. Voelt u zich, vanwege problemen met het zien, meer afhankelijk van anderen dan u zou willen? (heeft u bijvoorbeeld vaker hulp van anderen nodig dan u zou willen?)

□ nooit
□ zelden
□ soms
□ vaak
□ altijd

**Einde van de vragenlijst.**

Nogmaals hartelijk dank voor uw tijd en medewerking.

Dol VL, Fuermaier ABM, Roelofs TAJ, Vrijling ACL, Heutink J, Jansonius NM. The 6-Item Vision-Related Quality of Life and Limitations Questionnaire: Evaluation of Psychometric Properties. *Transl Vis Sci Technol*. 2024;13(3):5. doi:10.1167/tvst.13.3.5. CC BY-NC-SA 4.0
