## Supplementary S2 for "Discriminative capacity of the 6-item Vision-related Quality of life and Limitations questionnaire (VQL-6)"

*DISCLAIMER: The following questionnaire is an unvalidated English translation of the original Dutch questionnaire.*

### Instructions

**First, read the question and the response options.** Then please tick the correct response. It is important that you give your own opinion and complete the questions without consulting others.

**Only 1 response is possible per question.** Complete the entire questionnaire and please don't skip any questions! If you are unsure of how to answer a question, you may give an estimate.

**Did you make a mistake?** If you made a mistake it is not a problem. Just black out the wrong response entirely, and tick the correct response. Like so:

■ = wrong response

☒ = correct response

**Please answer all questions as if you were wearing your glasses or contact lenses (if any).**

The questions in this questionnaire relate to **your situation in the past month.**

Dol VL, Fuermaier ABM, Roelofs TAJ, Vrijling ACL, Heutink J, Jansonius NM. The 6-Item Vision-Related Quality of Life and Limitations Questionnaire: Evaluation of Psychometric Properties. *Transl Vis Sci Technol*. 2024;13(3):5. doi:10.1167/tvst.13.3.5. CC BY-NC-SA 4.0

1. How would you rate your health on a scale of 0 to 10?

*(0 = very poor, 10 = excellent)*

□ 0 □ 1 □ 2 □ 3 □ 4 □ 5 □ 6 □ 7 □ 8 □ 9 □ 10

2. How would you rate your quality of life on a scale of 0 to 10?

*(0 = very poor, 10 = excellent)*

□ 0 □ 1 □ 2 □ 3 □ 4 □ 5 □ 6 □ 7 □ 8 □ 9 □ 10

3. To what extent are you limited in your daily life because of problems with your vision? *(0 = no limitations, 10 = very limited)*

□ 0 □ 1 □ 2 □ 3 □ 4 □ 5 □ 6 □ 7 □ 8 □ 9 □ 10

4. Are you worried about the future because of problems with your vision?

□ never
□ rarely
□ sometimes
□ often
□ always

**See questions 5 and 6 on the next page**

Dol VL, Fuermaier ABM, Roelofs TAJ, Vrijling ACL, Heutink J, Jansonius NM. The 6-Item Vision-Related Quality of Life and Limitations Questionnaire: Evaluation of Psychometric Properties. *Transl Vis Sci Technol*. 2024;13(3):5. doi:10.1167/tvst.13.3.5. CC BY-NC-SA 4.0

5. Do you, because of problems with your vision, accomplish less than you would like or are limited in how long you can do things? (Think reading, working, household chores, going out or practicing hobbies)

- ☐ never
- ☐ rarely
- ☐ sometimes
- ☐ often
- ☐ always

6. Do you, because of problems with your vision, feel more dependent on others than you would like? (For example, do you need more help from others than you would like?)

- ☐ never
- ☐ rarely
- ☐ sometimes
- ☐ often
- ☐ always

**End of questionnaire.**

Thank you again for your time and cooperation.

Dol VL, Fuermaier ABM, Roelofs TAJ, Vrijling ACL, Heutink J, Jansonius NM. The 6-Item Vision-Related Quality of Life and Limitations Questionnaire: Evaluation of Psychometric Properties. *Transl Vis Sci Technol*. 2024;13(3):5. doi:10.1167/tvst.13.3.5. CC BY-NC-SA 4.0
